# PVT1 and MALAT1 Upregulation Correlates with Immune Checkpoints and Outcomes in African American Lung Cancer

**DOI:** 10.1101/2025.04.06.25325323

**Authors:** Lu Gao, Pushpa Dhilipkannah, Feng Jiang

## Abstract

**Background:** African Americans have the highest incidence and mortality rates of lung cancer among racial groups. Although therapies targeting the programmed cell death protein 1 (PD-1) and programmed death-ligand 1 (PD-L1) pathways have revolutionized treatment, their efficacy remains inconsistent, and adverse effects present significant challenges. Understanding the association between molecular changes and immune factors in the tumor immune microenvironment is critical for improving treatment outcomes and addressing racial disparities. Our previous research demonstrated that the upregulation of the long non-coding RNAs PVT1 and MALAT1 is associated with lung cancer in African Americans. This study investigates the relationship between dysregulated long non-coding RNAs, immune markers, and survival outcomes in a large cohort of lung cancer patients.

**Methods:** Tumor tissues were collected from 256 lung cancer patients, including 125 African Americans and 131 White Americans, who underwent lobectomy or pneumonectomy. PVT1 and MALAT1 expression levels were quantified using droplet digital polymerase chain reaction. Immune markers, including programmed death-ligand 1, programmed cell death protein 1, CD8^+^ T cell infiltration, CD69, interferon-gamma, and tumor necrosis factor-alpha, were analyzed through immunohistochemistry. Statistical analyses were performed to evaluate relationships between molecular changes, immune markers, and survival outcomes in this large cohort.

**Results:** Elevated PVT1 expression was associated with increased levels of programmed death-ligand 1 and programmed cell death protein 1, reduced CD69 expression, lower levels of interferon-gamma and tumor necrosis factor-alpha, advanced disease, and poorer survival outcomes in African American patients. MALAT1 upregulation was correlated with increased levels of programmed death-ligand 1 and programmed cell death protein 1, reduced immune effector activity, and worse survival outcomes in both African American and White American patients. Combined analysis of PVT1 and MALAT1 expression improved prognostic accuracy in African American lung cancer patients compared to individual assessments.

**Conclusions:** The dysregulation of PVT1 and MALAT1 is associated with immune evasion and advanced lung cancer among African Americans. PVT1 and MALAT1 may serve as prognostic biomarkers and therapeutic targets to improve the efficacy of immunotherapy for African American lung cancer patients. Leveraging these insights has the potential to reduce racial disparities in lung cancer outcomes and advance personalized treatment strategies.

**Key Points:**

- **Significant findings of the study**
  - Dysregulated PVT1 and MALAT1 are associated with immune evasion, advanced lung cancer, and poor survival in African Americans.
  - Combined analysis of PVT1 and MALAT1 enhances prognostic accuracy compared to individual assessments.
- **What this study adds**
  - Highlights PVT1 and MALAT1 as prognostic biomarkers and therapeutic targets in African American lung cancer patients.
  - Provides insights to improve immunotherapy efficacy and address racial disparities in lung cancer outcomes.

## Introduction

Non-small cell lung cancer (NSCLC) accounts for approximately 85% of all lung cancer cases and is the leading cause of cancer-related deaths among both men and women [1]. NSCLC is primarily divided into two main histological types: adenocarcinoma (AC) and squamous cell carcinoma (SCC) [1]. Significant disparities are evident in NSCLC incidence and mortality across different ethnic groups, with African American (AA) individuals experiencing higher prevalence and death rates [2]. Specifically, the annual lung cancer incidence is highest among AAs at 76.1 per 100,000, followed by White Americans (WAs) at 69.7 per 100,000, American Indians/Alaska Natives at 48.4 per 100,000, and Asian/Pacific Islanders at 38.4 per 100,000 [2]. The age-adjusted lung cancer mortality rate for AAs is approximately 46.6 per 100,000, which is significantly higher than that for WAs (34.3 per 100,000), American Indians/Alaska Natives (48.4 per 100,000), and Asian/Pacific Islanders (38.4 per 100,000) [2]. These disparities highlight the urgent need for targeted interventions and improved treatment strategies to address the higher mortality burden faced by AA lung cancer patients.

Immunotherapy targeting programmed death-1 (PD-1) and its ligand PD-L1 has significantly transformed cancer treatment by enhancing the capacity of the immune system to detect and destroy tumor cells [3]. These therapies have led to improved survival rates and durable responses in patients with NSCLC[3]. However, limitations such as variable response rates and immune-related adverse events necessitate new approaches to improve immunotherapy outcomes, particularly for AA populations who are disproportionately affected[3].

AA cancer patients may exhibit distinct molecular genetics, tumor biology, and immune profiles, potentially impacting their responses to various therapeutic interventions [4, 5]. Long non-coding RNAs (lncRNAs), which are over 200 nucleotides in length, are integral to various stages of tumor initiation, advancement, and metastasis [6]. In lung cancer specifically, lncRNAs function as essential regulatory molecules, influencing multiple pathways that drive tumorigenesis [6]. Our recent research revealed that AA lung cancer patients have higher plasma levels of the lncRNAs MALAT1 and PVT1 compared to cancer-free individuals, suggesting their potential as diagnostic biomarkers for lung cancer in AA individuals [7]. Furthermore, we found that dysregulation of MALAT1 may contribute to lung cancer disparities in AAs by modifying the tumor immune microenvironment through interactions with miR-206[7]. These findings suggest that the lncRNAs may play a role in immune evasion within the tumor immune microenvironment. This study aims to determine the association of dysregulated lncRNAs with immune factors in the tumor immune microenvironment and their impact on survival in AA lung cancer patients.

## Materials and Methods

### Patients and clinical specimens

This study was approved by the Institutional Review Board (IRB) of the University of Maryland (HP-00040666). Surgically resected lung tumor tissues and matched non-cancerous lung tissues were collected from 256 non-small cell lung cancer patients, comprising 125 AAs and 131 WAs, who underwent either lobectomy or pneumonectomy. Detailed demographic and clinical characteristics are presented in ***Table 1***. Histologically, 138 patients were diagnosed with AC and 118 with SCC. Staging distribution included 91 patients at stage I, 85 at stage II, and 80 at stages III-IV. Patients were followed from diagnosis until death or last follow-up. Overall Survival (OS) was used to evaluate the predictive potential of the genes for patient outcomes.

**Table 1:** Clinical and Demographic Characteristics of NSCLC Patients.

| Characteristics | NSCLC Patients (n = 256) |
| --- | --- |
| Age (Mean ± SD, years) | 65 (14.8) |
| Sex |  |
| - Female | 113 |
| - Male | 143 |
| Race |  |
| - AA | 125 |
| - WA | 131 |
| Smoking Pack-Years (Median) | 43 |
| Stage |  |
| - Stage I | 91 |
| - Stage II | 85 |
| - Stage III-IV | 80 |
| Histological Type |  |
| - AC | 138 |
| - SCC | 118 |

NSCLC, non-small cell lung cancer; AA, African Americans; WA, White Americans; AC, adenocarcinoma; SCC, squamous cell carcinoma; SD, standard deviation.

### Droplet Digital PCR (ddPCR)

PVT1 and MALAT1 were quantified using ddPCR as previously described with specific primer and probes [7]. In brief, RNA was reverse transcribed with the TaqMan miRNA RT Kit (Thermo Fisher Scientific, Waltham, MA). The ddPCR reactions were prepared by combining cDNA, Supermix, and TaqMan primer/probe mix, and subsequently processed in a QX100 Droplet Generator (Bio-Rad Laboratories, Hercules, CA), which generated over 10,000 droplets per well. Fluorescence detection was employed to accurately quantify lncRNA levels in each droplet. Target gene concentrations were calculated based on Poisson distribution, as detailed in prior studies [7, 8].

### Immunohistochemical (IHC)

To assess CD8^+^ T cells, PD-1, PD-L1, CD69, IFN-γ, and TNF-α, tissue specimens were fixed, paraffin-embedded, and sectioned. Sections underwent antigen retrieval, incubation with primary antibodies (Abcam. Waltham, MA), and secondary antibody treatment. Chromogen development and counterstaining were performed, followed by microscopy (Olympus. Center Valley, PA). For the quantitative analysis, five representative regions encompassing both cell and stroma areas were carefully selected. The positive cells were counted, and an average was calculated for each area. The resulting values for every hundred cells were classified according to the following scale: 0 (0–10% positive cells), 1 (11–25% positive cells), 2 (26– 50% positive cells), and 3 (more than 50% positive cells) [7]. Cells marked by a distinct, strong brown stain were manually counted using the AnalySIS Pro software (Olympus). The counting process was independently performed three times for each area by two staff, ensuring objectivity. All image analyses were conducted in a blind manner to avoid bias.

### Statistical Analysis

The Mann–Whitney U test was utilized for comparing continuous variables between groups, while the Chi-Square test was applied for categorical variables. Pearson’s correlation analysis assessed the linear relationships between lncRNA expression levels and immune factors, with Spearman’s rank correlation used if Pearson’s assumptions were not met. Kaplan–Meier survival curves were generated to compare survival outcomes between different lncRNA expression groups, with the log-rank test determining statistical significance. Multivariate Cox Proportional Hazards Regression Analyses were also used to evaluate the independent prognostic impact of lncRNA expression on survival outcomes. All statistical analyses were performed using SAS version 9.4, with a significance threshold set at p < 0.05.

## Results

### In AA lung cancer patients, elevated expressions of PVT1 and MALAT1 are associated with increased immune checkpoint activity and immune suppression

PVT1 expression was elevated in lung tumor tissues compared to non-cancerous lung tissues in AA lung cancer patients, but not in WA lung cancer patients (all p < 0.05) (Table 2). In both AA and WA lung cancer patients, MALAT1 expression was significantly higher in tumor tissues compared to non-cancerous lung tissues (Tables 2-3). Furthermore, MALAT1 expression was significantly higher in the tumor tissues of AA lung cancer patients than in WA patients (Tables 2-3). IHC analysis of CD8^+^ T cells, PD-1, PD-L1, CD69, IFN-γ, and TNF-α was successfully conducted on the tissue specimens. The expression levels of CD8^+^ T cells, PD-1, PD-L1, CD69, IFN-γ, and TNF-α varied across the tissue specimens, as illustrated in Figure 1. PD-L1 and PD-1 levels were significantly higher in tumor tissues than in non-cancerous lung tissues in both AA and WA patients (all p < 0.05) (Tables 2-3). In both populations, there was no significant change in the percentages of CD8^+^ T cells between tumor tissues and non-cancerous tissues (all p > 0.05). However, CD69 expression and levels of the pro-inflammatory cytokines IFN-γ and TNF-α were significantly reduced in tumor tissues relative to non-cancerous tissues in both AA and WA lung cancer patients (all p < 0.05) (Tables 2-3). No significant differences were observed in the levels of PD-L1, PD-1, CD8^+^ T cells, CD69 expression, IFN-γ, or TNF-α in tumor tissues between AA and WA lung cancer patients, nor between AC and SCC histological types (all p >0.05) (Tables 2-3).

**Table 2:** Expression Levels of lncRNAs and Immune Markers in AA and WA Lung Cancer Patients.

| Markers | Tumor Tissues<br>of AA Patients | Non-Cancerous<br>Tissues of AA<br>Patients | p-value (AA<br>Tumor vs<br>AA Non-Cancerous) | Tumor Tissues of<br>WA Patients | Non-Cancerous<br>Tissues of WA<br>Patients | p-value (WA<br>Tumor vs<br>WA Non-Cancerous) | p-value (AA<br>Tumor<br>vs WA Tumor) |
| --- | --- | --- | --- | --- | --- | --- | --- |
| MALAT1 Expression<br>(copies/μL) | 2.813 ± 0.495 | 1.502 ± 0.295 | 0.011 | 2.605 ± 0.403 | 1.405 ± 0.202 | 0.021 | 0.033 |
| PVT1 Expression<br>(copies/μL) | 2.705 ± 0.597 | 1.104 ± 0.302 | 0.012 | 1.003 ± 0.199 | 1.001 ± 0.201 | 0.946 | 0.031 |
| PD-L1 (% Positive Cells) | 27.481 ± 7.487 | 24.769 ± 4.823 | 0.048 | 36.951 ± 7.291 | 34.519 ± 4.496 | 0.046 | 0.321 |
| PD-1 (% Positive Cells) | 21.498 ± 6.476 | 18.032 ± 4.013 | 0.022 | 21.312 ± 6.231 | 17.792 ± 3.891 | 0.019 | 0.452 |
| CD8<sup>+</sup> T Cells (% Positive<br>Cells) | 15.024 ± 5.012 | 11.889 ± 4.682 | 0.152 | 14.782 ± 4.808 | 11.702 ± 4.482 | 0.121 | 0.603 |
| CD69 Expression (%<br>Positive Cells) | 9.013 ± 2.995 | 9.478 ± 3.599 | 0.041 | 8.223 ± 3.108 | 9.023 ± 3.513 | 0.346 | 0.023 |
| IFN-γ (% Positive Cells) | 10.012 ± 4.981 | 15.041 ± 4.789 | 0.033 | 9.195 ± 5.496 | 12.724 ± 4.881 | 0.027 | 0.759 |
| TNF-α (% Positive Cells) | 18.017 ± 6.002 | 23.021 ± 5.186 | 0.026 | 18.294 ± 5.793 | 22.813 ± 5.124 | 0.024 | 0.829 |

**Table 3:**
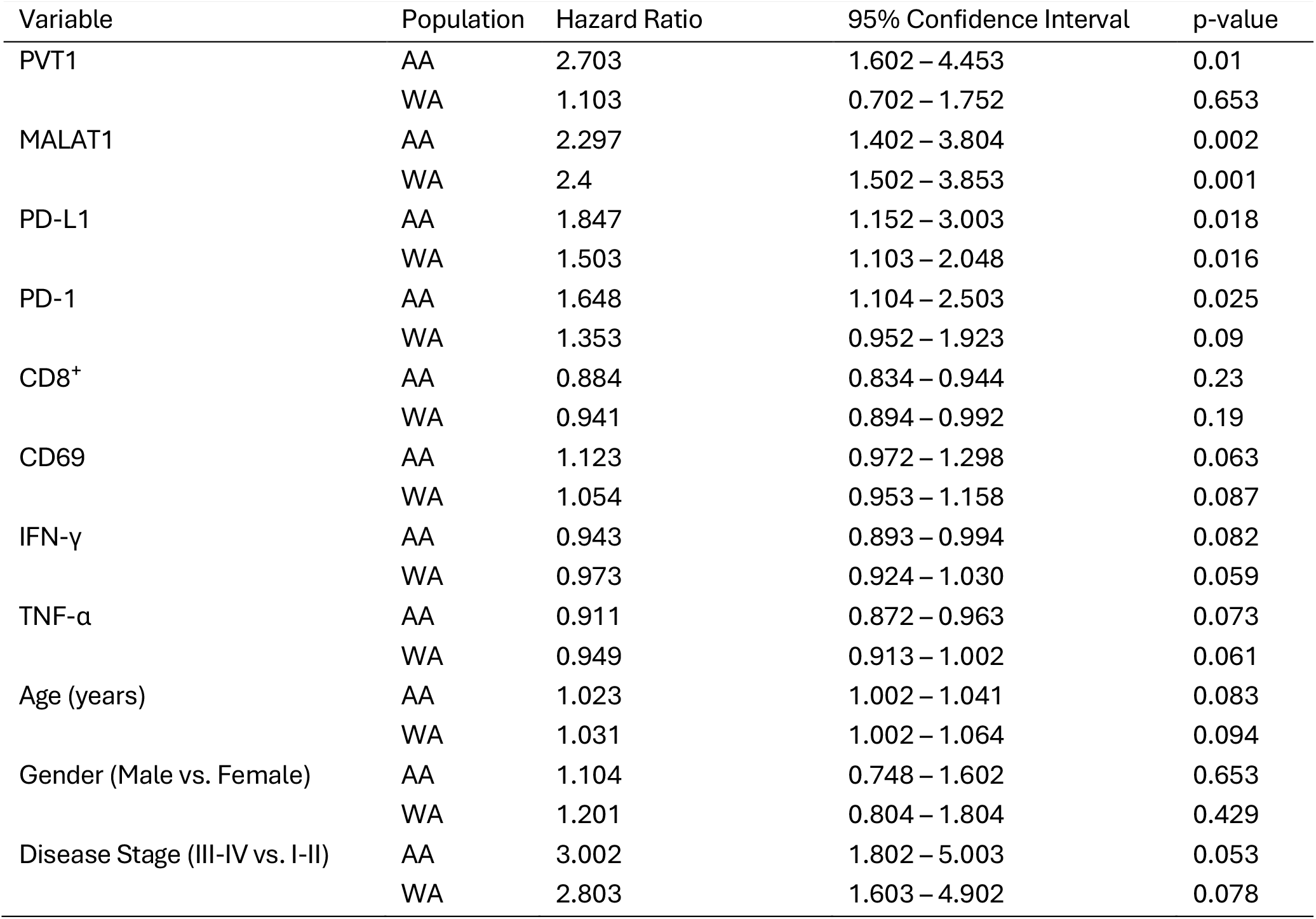
Multivariate Cox Proportional Hazards Regression Analysis of lncRNA Expression and Immune Factors, and Stags in AA and WA Lung Cancer Patients.

| Variable | Population | Hazard Ratio | 95% Confidence Interval | p-value |
| --- | --- | --- | --- | --- |
| PVT1 | AA | 2.703 | 1.602 – 4.453 | 0.01 |
|  | WA | 1.103 | 0.702 – 1.752 | 0.653 |
| MALAT1 | AA | 2.297 | 1.402 – 3.804 | 0.002 |
|  | WA | 2.4 | 1.502 – 3.853 | 0.001 |
| PD-L1 | AA | 1.847 | 1.152 – 3.003 | 0.018 |
|  | WA | 1.503 | 1.103 – 2.048 | 0.016 |
| PD-1 | AA | 1.648 | 1.104 – 2.503 | 0.025 |
|  | WA | 1.353 | 0.952 – 1.923 | 0.09 |
| CD8<sup>+</sup> | AA | 0.884 | 0.834 – 0.944 | 0.23 |
|  | WA | 0.941 | 0.894 – 0.992 | 0.19 |
| CD69 | AA | 1.123 | 0.972 – 1.298 | 0.063 |
|  | WA | 1.054 | 0.953 – 1.158 | 0.087 |
| IFN-γ | AA | 0.943 | 0.893 – 0.994 | 0.082 |
|  | WA | 0.973 | 0.924 – 1.030 | 0.059 |
| TNF-α | AA | 0.911 | 0.872 – 0.963 | 0.073 |
|  | WA | 0.949 | 0.913 – 1.002 | 0.061 |
| Age (years) | AA | 1.023 | 1.002 – 1.041 | 0.083 |
|  | WA | 1.031 | 1.002 – 1.064 | 0.094 |
| Gender (Male vs. Female) | AA | 1.104 | 0.748 – 1.602 | 0.653 |
|  | WA | 1.201 | 0.804 – 1.804 | 0.429 |
| Disease Stage (III-IV vs. I-II) | AA | 3.002 | 1.802 – 5.003 | 0.053 |
|  | WA | 2.803 | 1.603 – 4.902 | 0.078 |

**Figure 1.**
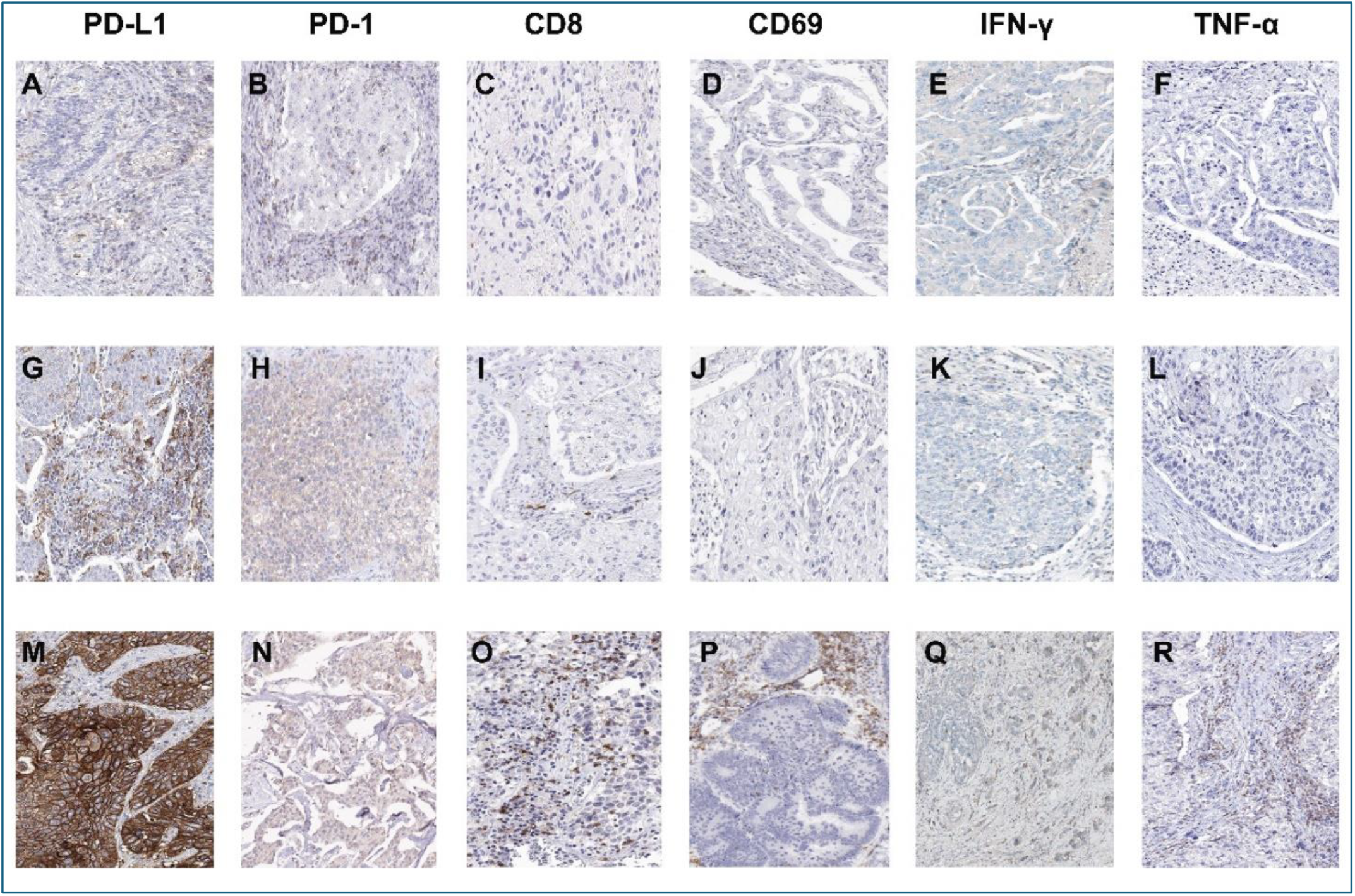
Immunohistochemical staining for PD-L1, PD-1, CD8, CD69, IFN-γ, and TNF-α in NSCLC tissues from AA patients. Panels A-F, N, Q, and R represent adenocarcinoma samples, while panels G-I, M, and P show squamous cell carcinoma samples. PD-L1 expression is evident on both tumor and immune cells, with 26–90% of cells showing positivity, whereas PD-1 is predominantly localized on T cells with 26–70% positivity. CD8^+^ T cells are observed infiltrating tumor regions, with 11–40% positive cells. CD69 expression is seen on T cells within tumor tissues, with 0–50% positivity. IFN-γ and TNF-α expression in T cells and macrophages is limited, with 0–45% positive cells. Images were captured at 200x magnification.

In the AA population, the elevated expression of the lncRNAs PVT1 and MALAT1 was positively correlated with increased levels of the immune checkpoint molecules PD-L1 and PD-1. Conversely, this elevated lncRNA expression was inversely associated with the reduced expression of the T cell activation marker CD69 and decreased levels of IFN-γ and TNF-α (all p <0.05) (Table 4). In WA lung cancer patients, although the correlation between MALAT1 expression and immune factors is weaker compared to that in AA lung cancer patients, elevated MALAT1 expression still showed positive correlations with PD-L1 and PD-1 levels and an inverse correlation with CD69 expression, IFN-γ, and TNF-α levels (Table 4). In contrast, PVT1 expression in WA lung cancer patients did not correlate with PD-L1, PD-1, CD8^+^ T cell levels, CD69 expression, IFN-γ, or TNF-α (all p >0.05) (Table 4). In In both AA and WA lung cancer patients, elevated levels of PD-L1 and PD-1 (all p < 0.05) are associated with advanced stages of NSCLC and poorer outcomes (Supplementary table 1). In contrast, reduced expressions of CD69, IFN-γ, and TNF-α is linked to improved survival and may provide benefits in advanced stages (all p < 0.05) (Supplementary table 1). CD8^+^ expression does not show significant associations with stage or survival (Supplementary table 1). Taken together, the observations imply that the upregulation of PVT1 and MALAT1 may contribute to immune suppression by enhancing immune checkpoint signaling and diminishing anti-tumor immune responses. Furthermore, PVT1 influences immune modulation in lung cancer patients within the AA population, while MALAT1 plays a crucial role in both AA and WA populations.

**Table 4:**
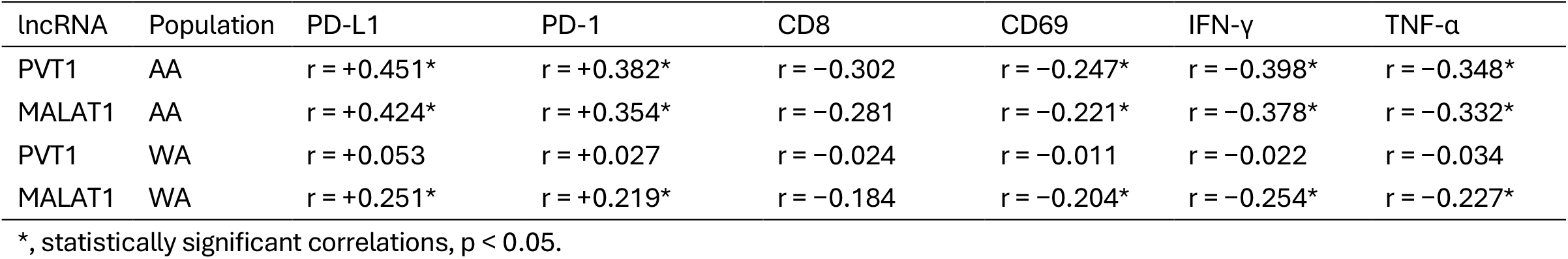
Pearson’s Correlation Coefficients between lncRNA Expression and Immune Factors in AA and WA Lung Cancer Patients.

| lncRNA | Population | PD-L1 | PD-1 | CD8 | CD69 | IFN-γ | TNF-α |
| --- | --- | --- | --- | --- | --- | --- | --- |
| PVT1 | AA | $r = +0.451^*$ | $r = +0.382^*$ | $r = -0.302$ | $r = -0.247^*$ | $r = -0.398^*$ | $r = -0.348^*$ |
| MALAT1 | AA | $r = +0.424^*$ | $r = +0.354^*$ | $r = -0.281$ | $r = -0.221^*$ | $r = -0.378^*$ | $r = -0.332^*$ |
| PVT1 | WA | $r = +0.053$ | $r = +0.027$ | $r = -0.024$ | $r = -0.011$ | $r = -0.022$ | $r = -0.034$ |
| MALAT1 | WA | $r = +0.251^*$ | $r = +0.219^*$ | $r = -0.184$ | $r = -0.204^*$ | $r = -0.254^*$ | $r = -0.227^*$ |

\*, statistically significant correlations, $p < 0.05$ .

### Prognostic significance of PVT1 and MALAT1 in survival among AA lung cancer patients

Elevated PVT1 expression levels are significantly associated with advanced disease stages and poorer overall survival outcomes in AA lung cancer patients (all p >0.05), but not in WA patients (Table 5), indicating its pivotal role in disease progression primarily within the AA population. Increased MALAT1 expression is significantly associated with advanced disease stages and poor survival outcomes in both AA and WA patients, indicating its broader impact across populations (all p >0.05). Furthermore, PVT1 and MALAT1 expression levels showed no significant associations with patient age, gender, or cancer histological type in either the AA or WA populations, indicating that their prognostic value in disease outcomes is independent of these demographic factors (Table 5).

**Table 5:** Associations of lncRNA Expression Levels with Immune Factors, Clinical Characteristics, and Overall Survival (OS) in AA and WA Lung Cancer Patients.

| Factor | Population | PVT1 (p-value) | MALAT1 (p-value) |
| --- | --- | --- | --- |
| PD-L1 | AA | 0.013 | 0.014 |
|  | WA | 0.853 | 0.012 |
| PD-1 | AA | 0.009 | 0.002 |
|  | WA | 0.917 | 0.018 |
| CD8<sup>+</sup> | AA | 0.078 | 0.094 |
|  | WA | 0.925 | 0.073 |
| CD69 | AA | 0.003 | 0.001 |
|  | WA | 0.947 | 0.027 |
| IFN-γ | AA | 0.002 | 0.003 |
|  | WA | 0.931 | 0.035 |
| TNF-α | AA | 0.004 | 0.003 |
|  | WA | 0.944 | 0.038 |
| Disease Stage | AA | 0.035 | 0.018 |
| - Stage I-II | AA |  |  |
| - Stage III-IV | AA |  |  |
|  | WA | 0.457 | 0.024 |
| - Stage I-II | WA |  |  |
| - Stage III-IV | WA |  |  |
| Overall Survival (OS) | AA | 0.017 | 0.011 |
| - Poor OS* | AA |  |  |
| - Good OS* | AA |  |  |
|  | WA | 0.307 | 0.014 |
| - Poor OS | WA |  |  |
| - Good OS | WA |  |  |
| Age (Mean ± SD, years) | AA | 0.653 | 0.657 |
|  | WA | 0.608 | 0.605 |
| Gender | AA | 0.752 | 0.748 |
| - Male | AA |  |  |
| - Female | AA |  |  |
|  | WA | 0.703 | 0.701 |
| - Male | WA |  |  |
| - Female | WA |  |  |
| Histological Type | AA | 0.217 | 0.402 |
| - Adenocarcinoma (AC) | AA |  |  |
| - Squamous Cell Carcinoma (SCC) | AA |  |  |
|  | WA | 0.603 | 0.507 |
| - Adenocarcinoma (AC) | WA |  |  |
| - Squamous Cell Carcinoma (SCC) | WA |  |  |

\*, Poor OS is defined as survival less than 60 months, while Good OS is survival of 60 months or more.

Interestingly, assessing the combined expression levels of PVT1 and MALAT1 provides higher prognostic accuracy for predicting survival in AA lung cancer patients compared to analyzing each lncRNA individually, highlighting their combined value as a strong prognostic indicator (Table 6) (Figure 2A). In contrast, for WA patients, high MALAT1 expression alone is associated with poorer survival (Table 6) (Figure 2B). The findings highlight the importance of integrating a panel of lncRNAs to enhance risk stratification and personalize treatment strategies for lung cancer in AAs.

**Table 6:** Multivariate Cox Proportional Hazards Regression Analysis of Combined PVT1 and MALAT1 Expression on Overall Survival in AA Lung Cancer Patients.

| Variable | Hazard Ratio | 95% Confidence Interval | p-value |
| --- | --- | --- | --- |
| PVT1 High Expression ($\geq 2$ copies/μL) | 2.723 | 1.602 – 4.457 | 0.013 |
| MALAT1 High Expression ($\geq 2$ copies/μL) | 2.305 | 1.398 – 3.793 | 0.026 |
| Combined PVT1 and MALAT1 High Expression ($\geq 2$ copies/μL for both) | 4.625 | 2.348 – 6.873 | 0.01 |

**Figure 2.**
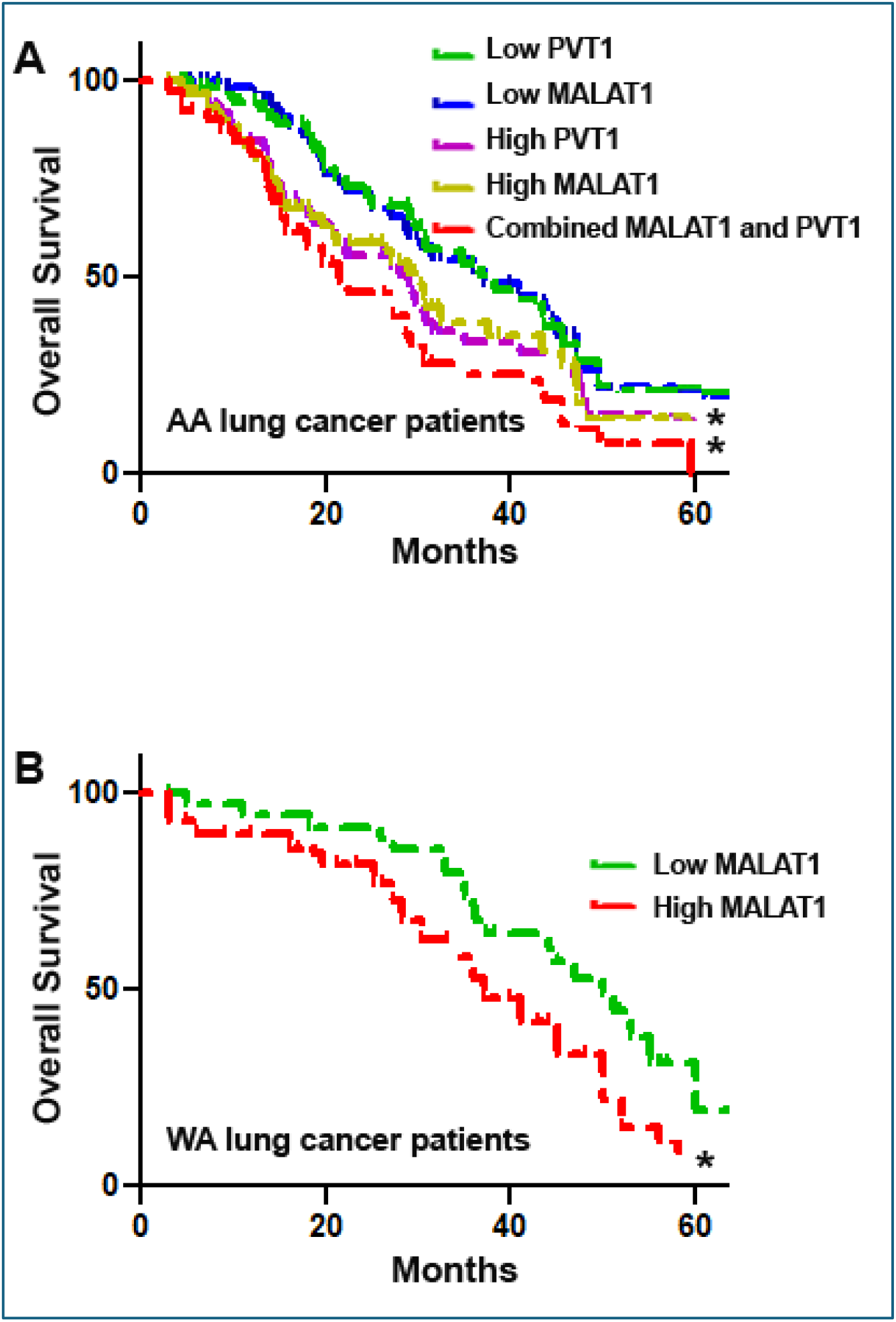
Kaplan-Meier Survival Curves Showing the Prognostic Value of PVT1 and MALAT1 Expression in Lung Cancer Patients. A: In AA patients, the high expression of either PVT1 (purple dash-dotted line) or MALAT1 (yellow dash-dotted line) alone is associated with significantly lower survival, while low expression of both lncRNAs (blue and green solid lines) indicates better outcomes. Combined high expression of PVT1 and MALAT1 (red dash-dotted line) shows a statistically significant survival difference compared to either PVT1 or MALAT1 alone (p < 0.05, log-rank test). B: In WA patients, high MALAT1 expression (red dashed line) is associated with poorer survival, whereas low expression of MALAT1 (green solid line) is linked to better survival outcomes (p < 0.05, log-rank test). *, p<0.05.

## Discussion

Previous studies have demonstrated the oncogenic roles of PVT1 and MALAT1 in various cancers, including lung cancer, by regulating gene expression, alternative splicing, and epigenetic modifications [9, 10]. PVT1 has been suggested to play a significant role in the development and progression of prostate cancer among AA patients [11]. Our recent research has shown that MALAT1 and PVT1 exhibit higher plasma levels in lung cancer patients from AA populations compared to WA patients [7]. Furthermore, we have demonstrated that MALAT1 interacts with miR-206, regulating monocyte chemoattractant protein-1 and macrophage activity, thereby contributing to tumorigenesis of AA lung cancer patients [7]. In the present study, we found that PVT1 and MALAT1 are significantly associated with increased expression of immune checkpoint molecules PD-L1 and PD-1, reduced expression of the activation marker CD69, and lower levels of pro-inflammatory cytokines IFN-γ and TNF-α in lung tumor tissues of AA patients. Although the percentages of CD8^+^ T cells in tumor tissues did not significantly change with increased PD-L1 and PD-1 expression, the upregulation of lncRNAs appears to contribute to T cell exhaustion. This is indicated by inverse correlations with CD69 expression and decreased levels of IFN-γ and TNF-α, potentially leading to functionally inhibited T cells [12]. Our findings indicate that the upegulation of PVT1 and MALAT1 may enhance immune checkpoint pathways, leading to a suppressed immune response within the tumor microenvironment. Consequently, dysregulation can promote tumor immune evasion and progression in lung tumorigenesis, particularly among AA patients.

While PD-1/PD-L1 immunotherapies have improved lung cancer survival, their variable efficacy and immune-related side effects underscore the need for optimized approaches, particularly for AA patients. Our findings on the association between elevated lncRNA expression and immune checkpoint activity might address this clinical need. Targeting these lncRNAs could enhance immunotherapy efficacy and reduce racial disparities in lung cancer outcomes, offering significant clinical implications for improving treatment strategies in AA patients.

MALAT1 expression correlates with poor clinical outcomes in both AA and WA lung cancer patients. However, PVT1 does not show a significant association with outcomes in the WA population, suggesting that PVT1 may play a more specific and critical role in disease progression within the AA population. This observation also has potential clinical implications, as integrating these two lncRNAs into clinical practice could improve prognostic assessments and support personalized treatment strategies for lung cancer in AA patients. Specifically, patients with high levels of both PVT1 and MALAT1 might benefit from combination therapies targeting these lncRNAs and immune checkpoints to restore T cell function and enhance anti-tumor immunity.

This study may include some limitations. The sample size, while sufficient to detect significant associations, may not fully represent the heterogeneity within AA and WA populations. Future research may focus on validating these findings in larger, multi-ethnic cohorts. Furthermore, environmental and lifestyle factors, notably smoking history, can significantly influence lncRNA expression and immune responses in lung cancer patients. Future research investigating the impact of environmental factors on lncRNA-related molecular pathways is crucial for developing comprehensive strategies to reduce lung cancer disparities. Additionally, the observational nature of the study limits the ability to establish causal relationships between lncRNA expressions and immune modulation. Functional studies are necessary to elucidate the direct mechanisms by which PVT1 and MALAT1 influence immune checkpoint pathways and T cell activity.

### Conclusions

Our research highlights the associations of PVT1 and MALAT1 with the tumor immune microenvironment and disease progression in NSCLC, with especially pronounced effects in AA patients. These two lncRNAs show potential as prognostic biomarkers and as targets for PD-L1-based immunotherapies, with the goal of improving clinical outcomes and addressing racial disparities in lung cancer prognosis and treatment.

## Data Availability

All data produced in the present study are available upon reasonable request to the authors

## Abbreviations

NSCLC: non-small cell lung cancer
AC: adenocarcinoma
SCC: squamous cell carcinoma
AA: African American
WA: White American
PD-1: programmed death-1
PD-L1: programmed death-1 ligand
lncRNA: long non-coding RNAs
MALAT1: Metastasis Associated Lung Adenocarcinoma Transcript 1
PVT1: Plasmacytoma Variant Translocation 1
IRB: Institutional Review Board
OS: Overall Survival
ddPCR: Droplet Digital PCR
IHC: Immunohistochemical

## Acknowledgements

The authors wish to thank the Biostatistics Shared Service at the University of Maryland Marlene and Stewart Greenebaum Cancer Center for their invaluable contribution in conducting the statistical analysis for this study.

## Authors’ Contributions

L. G and P.D performed experiments and wrote the main manuscript. F.J. developed the main project idea and conceptual framework. All authors reviewed the manuscript.

## Disclosure

The authors declare no conflict of interest

---

**Supplementary Table 1:** Associations of Immune Marker Expression Levels with NSCLC Stage, Survival, Age, Race, and Histological Type.

| Markers | Association with Advanced Stage (Stage III-IV vs I-II), p-value | Association with Survival, p-value | Association with Age, p-value | Association with Race, p-value | Association with Histology (AC vs SCC), p-value |
| --- | --- | --- | --- | --- | --- |
| PD-L1 | 0.014 | 0.009 | 0.647 | 0.578 | 0.602 |
| PD-1 | 0.017 | 0.012 | 0.725 | 0.608 | 0.553 |
| CD69 | 0.022 | 0.019 | 0.685 | 0.557 | 0.621 |
| IFN-γ | 0.019 | 0.016 | 0.703 | 0.618 | 0.583 |
| TNF-α | 0.025 | 0.021 | 0.662 | 0.595 | 0.635 |
| CD8 | 0.553 | 0.605 | 0.749 | 0.652 | 0.678 |

T-tests were used for comparisons across groups (stage, histological types, age, race). Cox proportional hazards models were used to assess associations with survival outcomes. A p-value < 0.05 was considered statistically significant.

## Notes

### Competing Interest Statement

The authors have declared no competing interest.

### Funding Statement

This study did not receive any funding

### Author Declarations

This study was approved by the Institutional Review Board (IRB) of the University of Maryland (HP-00040666).

